# Machine learning uncovers blood test patterns subphenotypes at hospital admission discerning increased 30-day ICU mortality rates in COVID-19 elderly patients

**DOI:** 10.1101/2022.05.10.22274889

**Authors:** Lexin Zhou, Nekane Romero-García, Rafael Badenes, Teresa García Morales, David Lora, Agustín Gómez de la Cámara, Francisco T García Ruiz, Juan M García-Gómez, Carlos Sáez

## Abstract

**Background:** Elderly patients with COVID-19 are among the most numerous populations being admitted in the ICU due to its high mortality rate and high comorbidity incidence. An early severity risk stratification at hospital admission could help optimize ICU usage towards those more vulnerable and critically ill patients.

**Methods:** Of 503 Spanish patients aged>64 years admitted in the ICU between 26 Feb and 02 Nov 2020 in two Spanish hospitals, we included 193 quality-controlled patients. The subphenotyping combined PCA and t-SNE dimensionality reduction methods to maximize non-linear correlation and reduce noise among age and full blood count tests (FBC) at hospital admission, followed by hierarchical clustering.

**Findings:** We identified five subphenotypes (Eld-ICU-COV19 clusters) with heterogeneous FBC patterns associated to significantly disparate 30-day ICU mortality rates ranging from 2% in a healthy cluster to 44% in a severe cluster, along three moderate clusters.

**Interpretations:** To our knowledge, this is the first study using age and FBC at hospital admission to early stratify the risk of death in ICU at 30 days in elderly patients. Our results provide guidance to comprehend the phenotypic classification and disparate severity patterns among elderly ICU patients with COVID-19, based only on age and FBC, that have the potential to establish target groups for early risk stratification or early triage systems to provide personalized treatments or aid the decision-making during resource allocation process for each target Eld-ICU-COV19 cluster, especially in those circumstances with resource scarcity problem.

**Funding:** FONDO SUPERA COVID-19 by CRUE-Santander Bank grant SUBCOVERWD-19.

Research in context

**Evidence before this study**
We searched on PubMed and Google Scholar using the search terms “COVID-19”, “SARS-CoV2”, “phenotypes” for research published between 2020 to 2022, with no language restriction, to detect any published study identifying and characterizing phenotypes among ICU COVID-19 patients. A previous COVID-19 phenotyping study found three phenotypes from hospitalized patients associated with significantly disparate 30-day mortality rates (ranging from 2·5 to 60·7%). However, it seems to become harder to find phenotypes with discriminative mortality rates among ICU COVID-19 patients. For example, we found one study that uncovered two phenotypes from 39 ICU COVID-19 patients based on biomarkers with 39% and 63% mortality rates, but such difference was not statistically significant. We also found another study with more success that uncovered two ICU COVID-19 phenotypes using two different trajectories with somehow disparate 28-day mortality rates of 27% versus 37% (Ventilatory ratio trajectories) and of 25% versus 39% (mechanical power trajectories).

**Added value of this study**
To our knowledge, this is the first study that uses age and laboratory results at hospital admission (i.e., before ICU admission) in elderly patients to early stratify, prior ICU admission, the risk of death in ICU at 30 days. We classified 193 patients with COVID-19, based on age and ten Full Blood Count (FBC) tests, into five subphenotypes (one healthy, three moderate, and one severe) that showed significantly disparate 30-day ICU mortality rates from 2% to 44%.

**Implications of all the available evidence**
Identifying, from elderly ICU patients with COVID-19 (Eld-ICU-COV19), subphenotypes could spur further investigation to analyze the potential differences in their underlying disease mechanisms, acquire better phenotypical understanding among Eld-ICU-COV19 toward better decision-making in distributing the limited resources (including both logistic and medical) as well as shedding light on tailoring personalized treatment for each specific target subgroup in future medical research and clinical trial.

## Introduction

Despite the promising vaccines and efforts, the coronavirus disease 2019 (COVID-19) outbreak is still outspread across the countries, exacerbating the burden on healthcare systems, depleting the limited resources, and taking away patients’ lives. Identifying distinct subphenotypes with disparate characterizations can help address such challenge. Disease subphenotyping plays a crucial role in personalized medicine, precision medicine, enrichment of clinical trials, better prognosis, and delivering tailored treatments to well-defined homogeneous patient subgroups, as demonstrated in recent years in many clinical settings^1,2^.

Patients with SARS-CoV-2 infection and associated COVID-19, frequently developed acute respiratory failure, requiring Intensive Care Unit (ICU) admission, with higher mortality rate and resource utilization –e.g., invasive ventilation, medication intake. Of which, elderly patients are among the most numerous populations being admitted in the ICU due to its high mortality rate and high comorbidity incidence^3,4^, where a reliable early risk stratification could help optimize ICU usage towards those more vulnerable and critically ill patients.

This study aims to uncover patterns in full-blood-count (FBC) tests measured at hospital admission in combination with the age that could early discern discrepancies in ICU mortality rates associated with distinct subphenotypes in Eld-ICU-COV19 patients.

## Methods

### Study design and population

This was a retrospective observational study at two Spanish hospitals, Hospital Universitario 12 de Octubre de Madrid and Hospital Clínico Universitario de Valencia. The data were available between 26 February 2020 and 02 November 2020. Patients with COVID-19 at hospital admission with a posterior ICU admission were eligible. This resulted in an initial set of 503 Spanish Eld-ICU-COV19 aged>64 years. The use of data was approved by the Ethical Committees of the two hospitals and the Universitat Politècnica de València. STROBE recommendations were followed (appendix 1).

The studied variables included patient demographics, FBC laboratory tests, and clinical outcomes. A data quality control was applied to exclude cases with missing data, as described next.

### Data preprocessing and Study measures

The study measures include for each patient the age, sex, date of hospitalization, date of ICU admission, date of death (if any), and ten FBC variables including Leukocytes (*K/µL*), Neutrophils (*%*), Eosinophils (*%*), Basophils (*%*), Monocytes (*%*), Lymphocytes (*%*), Erythrocytes (*Mill/µL*), Hemoglobin (*g/dL*), Hematocrit (*%*), and Platelets (*K/µL*). All these variables were obtained and measured at the time of hospitalization.

Also, we derived three variables by using the date of hospitalization, the date of ICU admission, and the date of death: (1) ADM2ICU, indicates how many days it took for a hospitalized patient to be admitted to ICU; (2) Mortality7_ICU, indicates whether a patient suffered death within seven days after being admitted to ICU; (3) Mortality30_ICU, indicates whether a patient suffered death within thirty days after being admitted to ICU.

We excluded patients who showed any missing data in the studied variables. Specifically, all the patients who showed any missing data had at least two variables with missing data discouraging the use of variable imputation methods since including few variables, and thus preferring the inclusion of a comprehensive set of individuals for the sake of robustness and reliability. Besides, we performed a principal components analysis (PCA)^5^ for both exploratory and evaluation purposes, where we found no outliers or further data quality issues. Hence, no patient was excluded in this second phase. This finally resulted in 193 quality-controlled patients. This selection process is summarized in the CONSORT flowchart in Figure 1.

**Figure 1.**
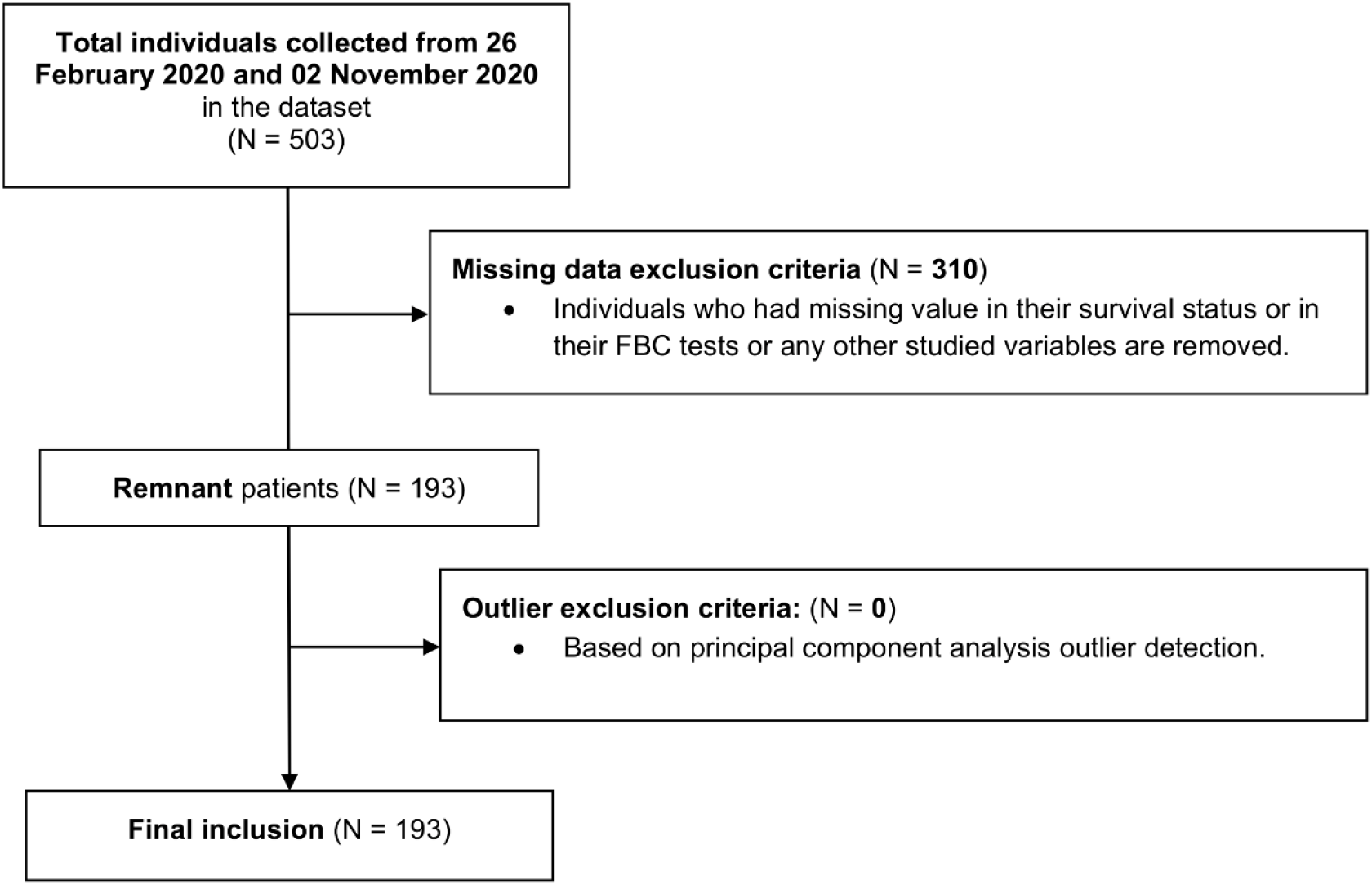
CONSORT flowchart about the inclusion/exclusion criteria of the final sample.

### Subphenotype clustering

The Machine Learning pipeline in this study first applied a non-linear dimensionality reduction of the variables using t-distributed stochastic neighbor embedding (t-SNE)^6^, fed by the normalized input variables using PCA as a robust t-SNE initialization method^7,8^. The t-SNE parameters were 5000 maximum iterations and a perplexity value of N^1/2^, where N is the number of patients. The t-SNE algorithm maximizes non-linear similarity among input variables, reducing noises, keeping distances between pairs of points while also preserving both local and global information –i.e., points that are close to one another in the high-dimensional dataset, will tend to be close to one another in the low dimension– after embedding the data into 3-dimensional subspace that also facilitates visualization.

We then performed a hierarchical clustering on the t-SNE output using Euclidean distance. The number of clinically relevant subphenotypes was chosen by combining two clustering quality metrics, namely the Silhouette Coefficient and Elbow Coefficient^9,10^, and validated with an exploratory analysis.

### Statistical Analysis

When analyzing the overall characteristics of each cluster, data are presented as mean accompanied by Standard Deviation (SD), and absolute number accompanied by percentage for numeric variables and categorical variables, respectively. Numeric variables’ means between clusters were compared using Tukey’s honest significance difference test (Tukey HSD)^11,12^ with 95% confidence level to verify the significant difference, i.e., verify which means amongst a set of means differ from the rest. Odds Ratio (OR) with a 95% Confidence Interval (CI) as well as the corresponding P-value using Fisher’s exact test^13^ were computed for comparing categorical variables between clusters. All analyses were done on R Studio, version 1.3.1073, using R, version 4.0.2.

### Role of the funding source

The funder of the study had no role in study design, data collection, data analysis, data interpretation, or writing of the report.

## Results

Between 26 February 2020 and 02 November 2020 503 elderly Covid 19 patients were admitted to the ICU of the two Spanish hospitals. Of the 503 patients, 310 were not screened because of the high missing data proportion. Among the 193 Eld-ICU-COV19 patients, 77 (39·9%) accounted for female, with a mean age of 76·26 years (SD 8·62), a mean ADM2ICU of 1·84 (SD 3·95) days, and a 30-day ICU mortality rate of 22% (43 patients), as described in Table 1.

**Table 1.**
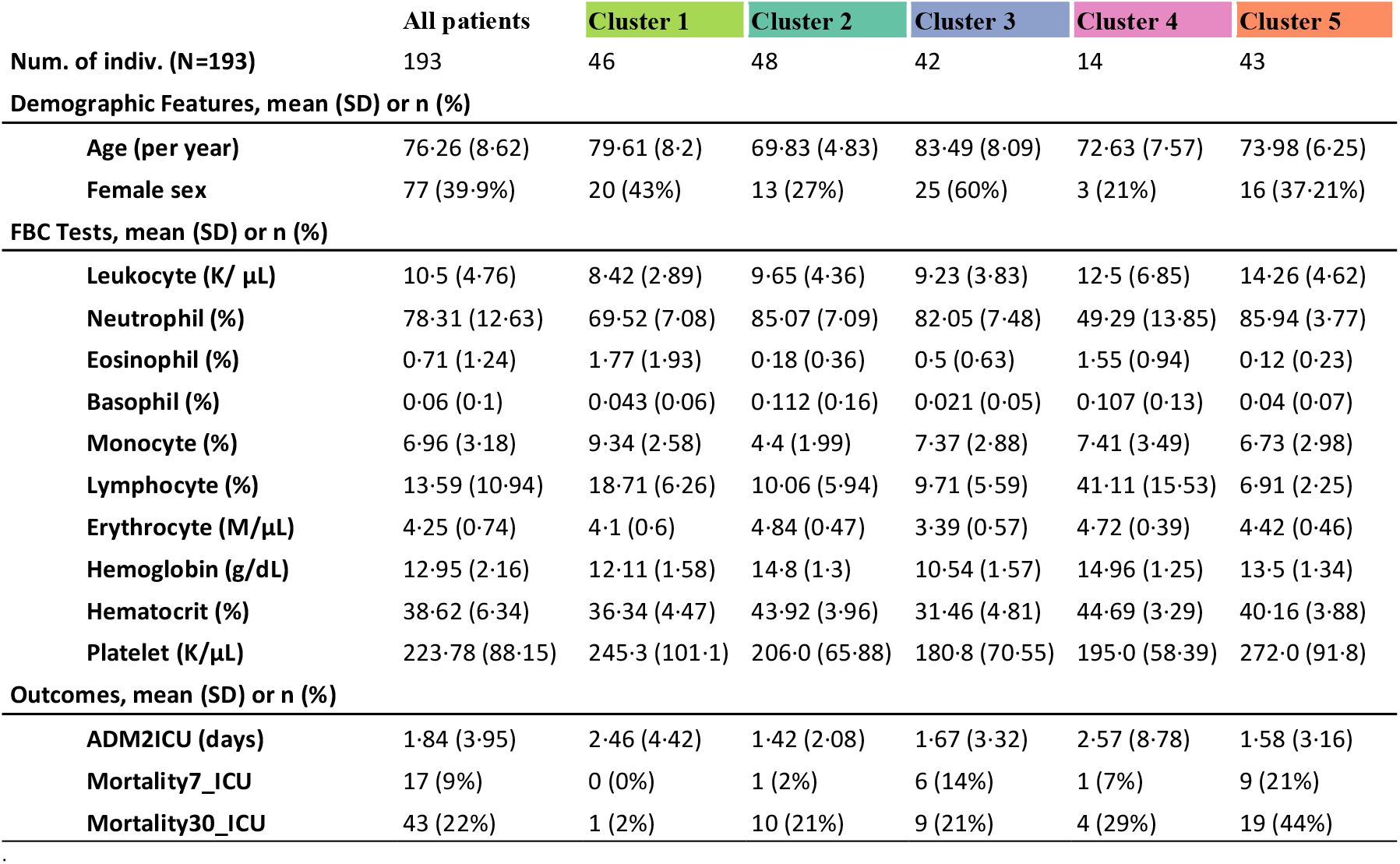
Main features of the 5 resultant Eld-ICU-COV19 clusters with 95% CI. Spain, February 26–02 November, 2020.

After analyzing the Silhouette Coefficient and Elbow Coefficient (Appendix 2), we identified five subphenotypes with heterogeneous inter-clustering age and FBC patterns associated with significantly disparate 30-day ICU mortality rates ranging from 2% to 44%. Each cluster counted with approximately 42-48 patients, except cluster 4 that only has 14 patients (Table 1). Figure 2 provides a visually effective comparison of the differences in normalized average FBC results between the five clusters and individual patterns for each cluster.

**Figure 2.**
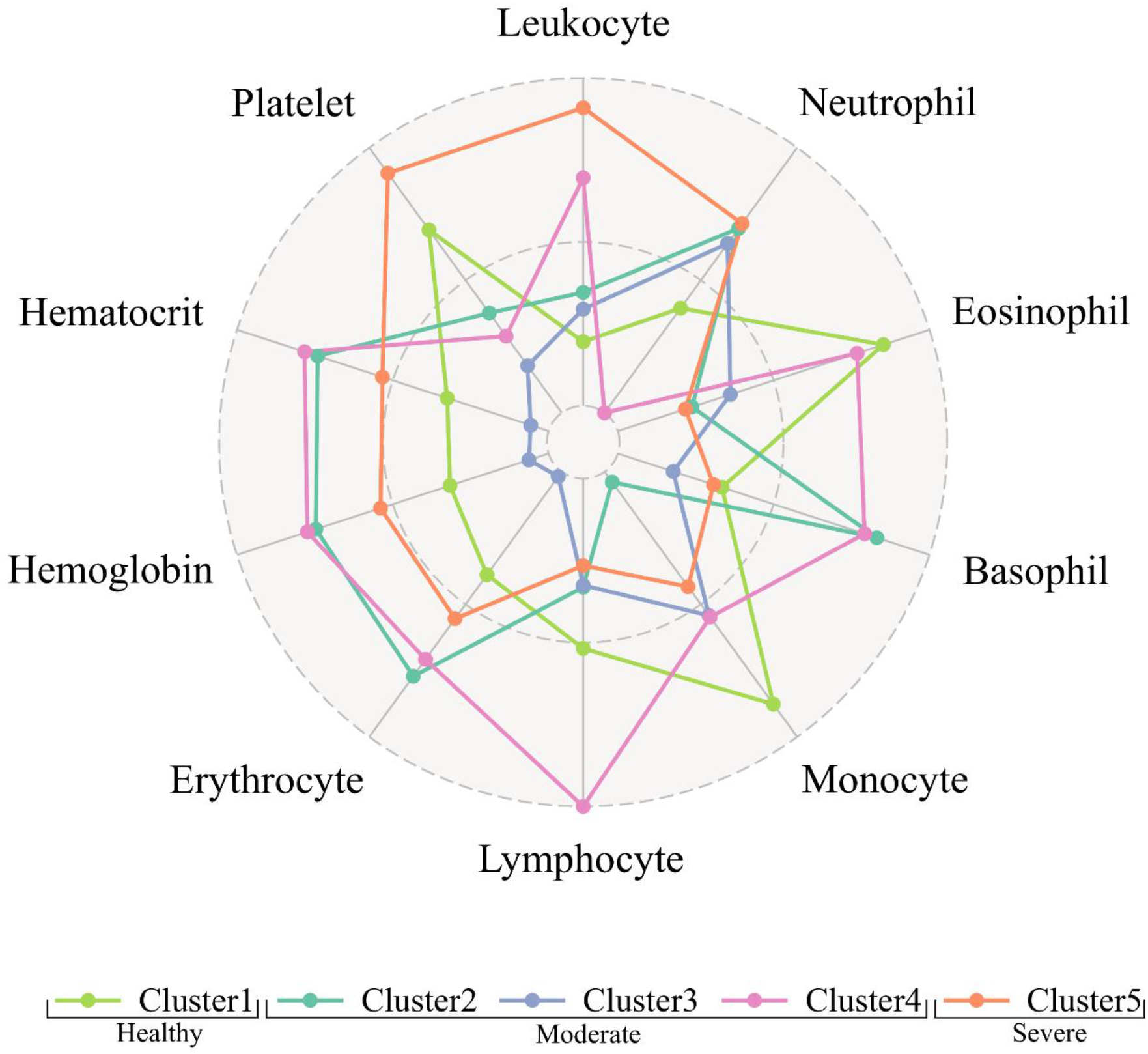
Differences in standardised average blood cells counts between the five Eld-ICU-COV19 subphenotypes. See Table 1 for the original values. Spain, February 26–02 November, 2020.

Figure 3 and Figure 4 show, respectively, the inter-clustering significant difference between the numeric variables (age, FBCs, and ADM2ICU), and the ICU mortality within 30 days using OR alongside the corresponding p-value. Appendix 3 describes the statistical difference between the variables sex and Mortality7_ICU.

**Figure 3.**
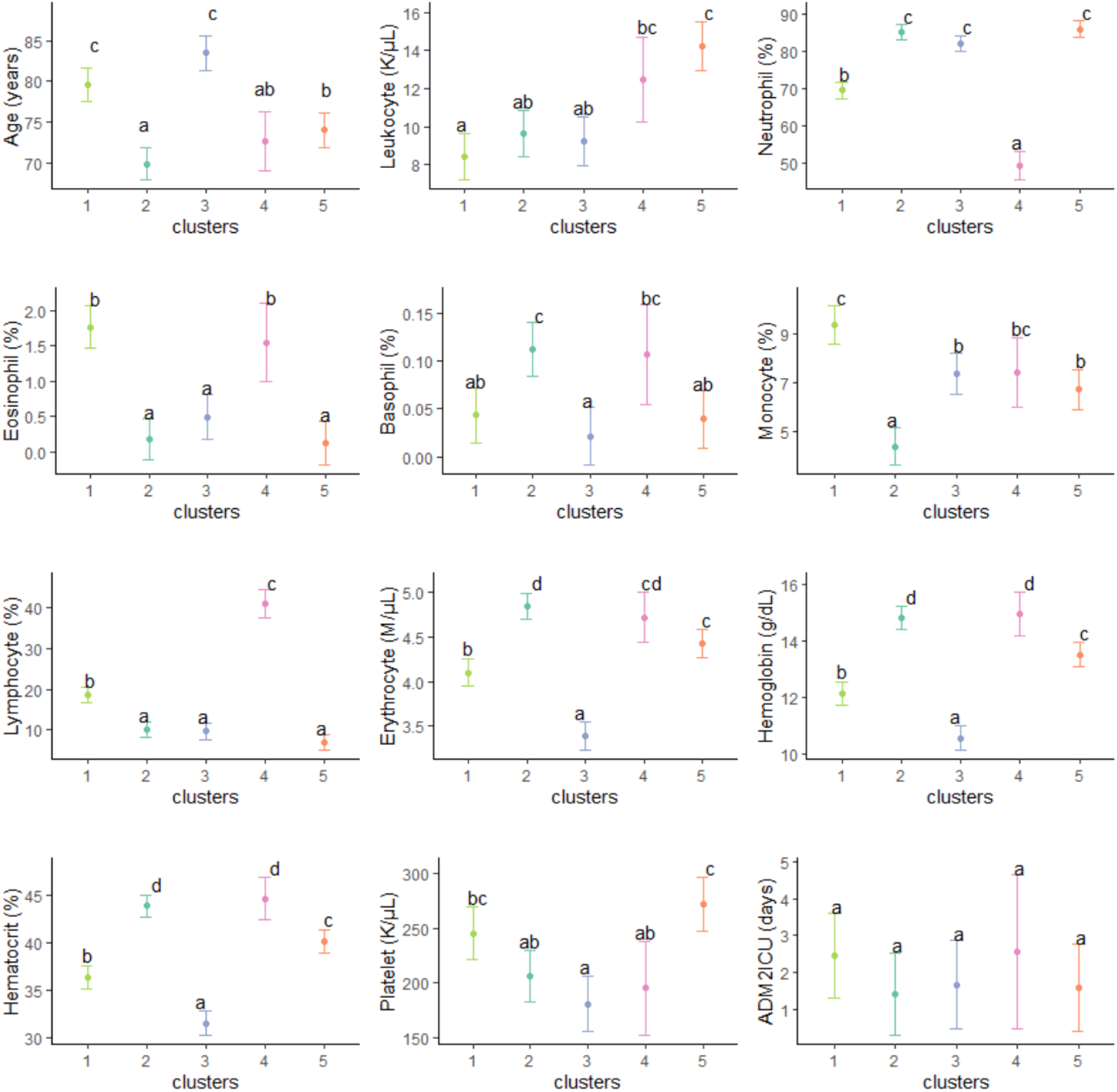
Tukey’s honest significance difference test with a 95% confidence interval for the studied numeric variables. Y-axis represents each variable, and the X-axis represents the cluster IDs. If two intervals overlap, they will have the same letter, such as a, b, c, and/or d, at the top of the interval.

**Figure 4.**
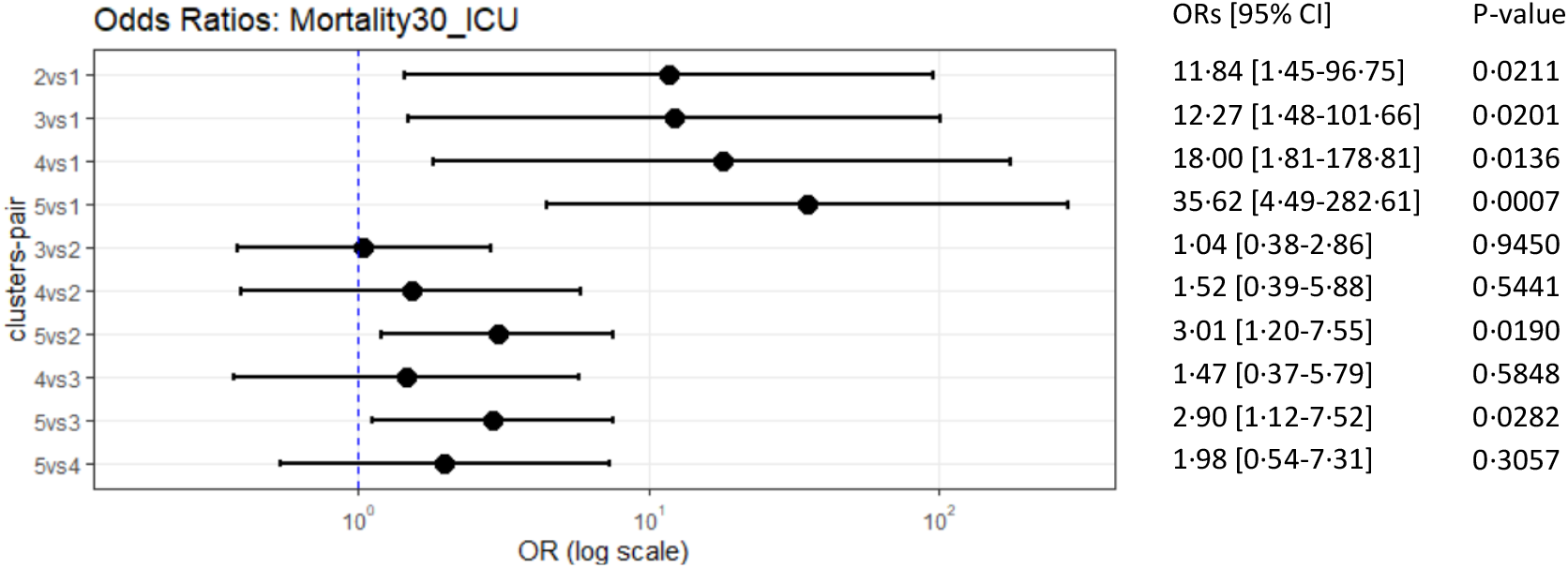
Odds Ratios with 95% CI and the corresponding p-value using Fisher’s exact test for the ICU mortality within 30 days. The Y-axis represents the clusters-pair that it compares. The X-axis represents the OR on the log-10 scale.

From the five subphenotypes, we found one ‘healthy’ subphenotype –cluster 1– that showed one case of (2%) 30-day ICU death incidence; characterized by relatively moderate count of leukocytes (8·42K/µL [SD=2·89]) and eosinophils (1·77% [SD=1·93]), the highest monocytes count (9.34% [SD=2·58]), low hemoglobin (12·11g/dL (SD=1·58)) and hematocrit counts (36·34% [SD=4·47]), and little count of erythrocytes (4,1M*/µL* [SD=0·6]).

One severe subphenotype –cluster 5– showed an increased 30-day ICU mortality rate of 44% (OR 35.63 [4.49-282.62; 95%CI] compared with the ‘healthy’ subphenotype; p=0.0007); characterized by abnormal high count of leukocytes (14·26K/µL [SD=4·62]) and neutrophils (85·94% [SD=3·77]), lacking monocytes (6·73% [SD=2·98]) and lymphocytes (6·91% [SD=2·25]), the lowest eosinophil (0·12% [SD=0·23]) and the highest platelet count (272K/µL [SD=91·8]).

Three moderate subphenotypes –cluster 2, 3, 4– with relatively higher 30-day ICU mortality rates (20%-28%) compared with the ‘healthy’ subphenotype –cluster 1– (p<=0.0211, Figure 4C). Cluster 2 is similar to cluster 5 with non-significant difference in terms of lacking lymphocyte (10·06% [SD=5·94]), lacking eosinophil (0·18% [SD=0·36]), and abundant neutrophil counts (85·07% [SD=7·09]), but differentiated by its lowest mean age (69·83years [SD=4·83]), a predominant male rate (13 [27%]), and the abundant count of red blood cells –erythrocyte (4·84M/µL [SD=0·47]), hemoglobin (14·8g/dL [SD=1·3]), and hematocrit (43·92% [SD=3·96])– (Figure 3).

Similar to cluster 2, cluster 3 shows a normal leukocyte count (9·23K/µL [SD=3·83]), little counts of lymphocyte (9·71K/µL [SD=5·59]) and eosinophil (0·5% [SD=0·63]), and abundant neutrophil count (82·05K/µL [SD=7·48]). However, it differentiates from the former by the highest mean age (83·49years [SD=8·09]), the highest female rate (25 [60%]), the lacking counts of red blood cells –erythrocyte (3·39M/µL [SD=0·57]), hemoglobin (10·54M/µL [SD=1·57]) and hematocrit (31·46% [SD=4·81])– and the lowest platelet count (180·8K/µL [SD=70·55]).

Cluster 4 is characterized by the lowest neutrophil (49·29% [SD=13·85]) and the highest lymphocyte counts (41·11% [SD=15·53]), with a noticeable significant difference among the five (Figure 3). Noteworthy, albeit that cluster 4 has a moderate 30-day ICU mortality rate of 28%, there is no statistically significant difference when we compare cluster 5 (the ‘severe’ one) versus cluster 4 in terms of 7-day ICU mortality (OR 3·44 [0·4-29·92; 95%CI]; p=0·2627) and 30-day ICU mortality (OR 1·98 [0·54-7·31; 95% CI]; p=0·3057).

## Discussion

This large multicenter retrospective study, has identified five Eld-ICU-COV19 subphenotypes with discriminative FBC patterns alongside age. Noteworthy, subphenotypes 1 and 5, with an ICU 30-day mortality rate of 2% to 44%, respectively, provide new insights on COVID-19 severity phenotypes from an early immunological presentation at hospital admission in elderly patients.

The phenotypic presentation of Eld-ICU-COV19 COVID-19 patients is not fully understood yet. Former studies used mechanisms for general COVID-19 patients profiling for this purpose. For instance, Sudre et al. reported non-hospitalized COVID-19 patient phenotypes based only on self-declaration of symptoms using an app^14^. Gutiérrez-Gutiérrez et al. used clinical data to uncover COVID-19 phenotypes, including hospitalized patients of all ages, and successfully found three phenotypes associated with significantly disparate 30-day mortality rates^15^. However, since ICU COVID-19 patients themselves are prone to a higher incidence of death, it seems to become harder to discern phenotypic patterns that discern different mortality levels associated with distinct ICU subphenotypes. For example, Sinha et al. used biomarkers and found two phenotypes from 39 ICU COVID-19 patients with 39% and 63% mortality rates, but without statistical difference^16^. A large cohort study Bos X et al. found two ICU COVID-19 phenotypes demonstrated certain success using two different trajectories with somehow disparate 28-day mortality rates of 27% versus 37% (Ventilatory ratio trajectories) and of 25% versus 39% (mechanical power trajectories)^17^ but may still not be sufficient to discriminate more disparate mortality rates associated with different COVID-19 phenotypes. In addition, these phenotyping analyses primarily focused on using variables obtained after ICU admission rather than leveraging the available information prior to ICU admission, which impedes the possibility of estimating the expected outcome for each phenotype prior to ICU admission.

Thus, subphenotying COVID-19 patients with ICU admission that display discriminative patterns associated with different mortality levels is crucial. In this report, we focused on elderly patients because they are within the most numerous populations being admitted in the ICU. A reliable early risk stratification could help optimize ICU usage towards those more vulnerable and critically ill patients. As a result, we identified five Eld-ICU-COV19 subphenotypes. The findings of this preliminary report of 193 Eld-ICU-COV19 patients suggest the patient’s FBC result can display discriminative patterns associated with disparate 30-day ICU mortality rates.

In our results, white blood cells seem to be most relevant factors that differentiate the cluster 1–the heathy cluster– and the cluster 5 –the severe cluster: the healthy cluster shows white cell counts which fall within normal boundaries, reflecting a physiological immune response. In contrast, the remarkable increase in total leukocytes, as well as neutrophils percentage, and the decrease in lymphocytes, all of which define the severe cluster, depict dysregulated response to infection and have previously been correlated to higher mortality^18,19,20^. In fact, the neutrophil-to-lymphocyte ratio (NLR) is probably the most frequently cited severity predictor^21^. Similarly, neutrophil to monocyte ratio, which is highest in the healthy cluster, seems to be another potential candidate in recent studies^18^. Even though eosinophils appear to have a limited role in this kind of immune response, previous studies found eosinophils percentage to significantly decrease in SARS-CoV-19 compared to other causes of pneumonia, and its recovery to be parallel to CT-scan improvement and therefore eventually in patient groups with higher recovery rates^22^. In addition, platelet count is within limits in all groups, but highest in the severe cluster, which may be explained by the role of platelets as acute phase reagents and therefore reflect a more intense immune response. It is important to highlight that most of these FBC were obtained during hospitalization, and very few studies have proven such correlations may already be present at admission.

The importance of NLR is remarkable in the two moderate clusters 2 and 3, whose NLR is similar to the severe cluster. However, in contrast to the seve cluster the former two clusters 2 and 3 show normal total leukocytes. The low-normal leukocyte count in clusters 2 and 3 might be explained by a previous cellular immunosuppression status, such as AIDS or immunosuppressive drugs affecting lymphocytes.

Regarding red blood cells, all clusters except from cluster 3 show in-range values of red blood cells and haemoglobin. This could be related to the oldest age of this group, age being a well-established risk factor for anemia. Interestingly, cluster 3 also shows the lowest number of platelets, which is opposite to what is expected in iron-deficiency anemia and could point to some underlying disease, such as CKF. Anemia being an aggravating condition for most diseases, it has also been suggested as a risk factor for severe outcomes in SARS-CoV-19 infection^18,20^ and could contribute to higher mortality in cluster 3.

Regarding the importance of demographic features, previous studies found age and gender as key factors in explaining the higher incidence of death^23,24,25^. However, we found that the roles of age and gender are not so straightforward in discriminating the associated 30-day ICU mortality in Eld-ICU-COV19 patients since the healthy cluster and the severe cluster do not have the highest/lowest mean age or male/female ratio, and also their Tukey HSD 95% CIs overlap with other moderate clusters. For instance, the mean age of the healthy cluster is not significantly different from the mean age of cluster 3 –a moderate cluster– and the mean age of the severe cluster is not significantly different from the mean age of cluster 4 –another moderate cluster. This suggests that FBC tests outweigh the age and gender in discriminating the increased 30-day ICU mortality among the five Eld-ICU-COV19 subphenotypes, possibly because here we only focused on a particular COVID-infected age group (elderly) admitted in ICU.

This study has several strengths. This is the first study, to our knowledge, that uses age and blood tests at hospital admission in elderly COVID-19 patients. The sole use of the age and blood tests obtained prior to ICU admission has the utility of early stratifying the risk of death in ICU at 30 days based on objective signs, and not requiring to acquire any further complex variables or costly information of our patients; meaning that the expected outcome of the patients from each target group can be inexpensively estimated beforehand if these patients are admitted into ICU. In addition, focusing on subphenotyping patients from a particular age group –elderly in our case– generally helps uncover more specific detailed subphenotypes and prevents overgeneralization^26,27^, which may have more pragmatical potential to shed light on tailoring personalized treatment for each specific target subgroup in future medical research and clinical trial.

One limitation of this study is its sample size. The small number of patients in some subphenotypes may make the comparative statistics sometimes difficult to interpret (e.g., cluster 4 only has 14 patients). In addition, our data are from two hospitals, which favors generalization, although future studies from third hospitals may benefit as external validations.

In summary, by using Machine Learning we identified five Eld-ICU-COV19 subphenotypes with discriminative FBC patterns alongside age. Of which, we found one healthy cluster –where nearly all patients survived within 30-day after ICU admission, one severe cluster –where nearly half of the patients lost their life within 30-day after ICU admission– and three moderate clusters –whose 30-day ICU mortality is similar to the populational level– with one that has the potential of being categorized into the severe category. Our results can provide guidance to comprehend the phenotypic classification and disparate severity patterns among elderly ICU patients with COVID-19, based only on age and FBC tests, that have the potential to establish target groups for an early risk stratification prior to ICU admission or early triage systems to provide personalized treatments or aid the decision-making during resource allocation process for each target Eld-ICU-COV19 group, especially in those circumstances with resource scarcity problem.

## Data Availability

All data used in the present study is not publicly available. Any expression of interest must be sent upon reasonable request to the authors.

## Abbreviations

Eld-ICU-COV19: Elderly ICU patients with COVID-19
ICU: Intensive Care Unit
WBC: White Blood Cell
RBC: Red Blood Cell
OR: Odds Ratio
CI: Confidence Interval
FBC: Full Blood Count
PCA: Principal Component Analysis
t-SNE: t-distributed Stochastic Neighbor Embedding

## Contributors

LZ, NRG, RB, AGC, JMGG, FTGR, and CS conceived this study. TGM and NR acquired the data. LZ and CS processed and analyzed the data and performed the statistical analysis. LZ, NRG, RB, AGC, JMGG, and CS were involved in the analysis or interpretation of the data. LZ, NR, and CS led the writing of the paper. LZ and FTGR drafted the figures. All authors revised the manuscript critically, approved the final manuscript, and are accountable for the accuracy and integrity of the manuscript.

## Appendix

## Appendix 1: STROBE Statement—Checklist

|  | Item No | Recommendation | Section or page in manuscript |
| --- | --- | --- | --- |
| Title and abstract | 1 | (a) Indicate the study’s design with a commonly used term in the title or the abstract | Title |
|  |  | (b) Provide in the abstract an informative and balanced summary of what was done and what was found | Abstract |
| Introduction |  |  |  |
| Background/rationale | 2 | Explain the scientific background and rationale for the investigation being reported | Introduction |
| Objectives | 3 | State specific objectives, including any prespecified hypotheses | Introduction, last paragraph |
| Methods |  |  |  |
| Study design | 4 | Present key elements of study design early in the paper | Methods |
| Setting | 5 | Describe the setting, locations, and relevant dates, including periods of recruitment, exposure, follow-up, and data collection | Methods |
| Participants | 6 | (a) Give the eligibility criteria, and the sources and methods of selection of participants | Methods |
| Variables | 7 | Clearly define all outcomes, exposures, predictors, potential confounders, and effect modifiers. Give diagnostic criteria, if applicable | Methods |
| Data sources/measurement | 8* | For each variable of interest, give sources of data and details of methods of assessment (measurement). Describe comparability of assessment methods if there is more than one group | Methods |
| Bias | 9 | Describe any efforts to address potential sources of bias | Discussion (paragraph discussion limitations) |
| Study size | 10 | Explain how the study size was arrived at | Methods |
| Quantitative variables | 11 | Explain how quantitative variables were handled in the analyses. If applicable, describe which groupings were chosen and why | Methods |
| Statistical methods | 12 | (a) Describe all statistical methods, including those used to control for confounding | Methods |
|  |  | (b) Describe any methods used to examine subgroups and interactions | Methods |
|  |  | (c) Explain how missing data were addressed | Methods |
|  |  | (d) If applicable, describe analytical methods taking account of sampling strategy | Not Applicable |
|  |  | (e) Describe any sensitivity analyses | Not Applicable |
| **Results** |  |  |  |
| Participants | 13* | (a) Report numbers of individuals at each stage of study—eg numbers potentially eligible, examined for eligibility, confirmed eligible, included in the study, completing follow-up, and analysed | Methods |
|  |  | (b) Give reasons for non-participation at each stage | Methods |
|  |  | (c) Consider use of a flow diagram | Methods |
| Descriptive data | 14* | (a) Give characteristics of study participants (eg demographic, clinical, social) and information on exposures and potential confounders | Table 1 |
|  |  | (b) Indicate number of participants with missing data for each variable of interest | There is no missing data in the studied sample |
| Outcome data | 15* | Report numbers of outcome events or summary measures | Methods and Results |
| Main results | 16 | (a) Give unadjusted estimates and, if applicable, confounder-adjusted estimates and their precision (eg, 95% confidence interval). Make clear which confounders were adjusted for and why they were included | Methods and Results |
|  |  | (b) Report category boundaries when continuous variables were categorized | Not applicable |
|  |  | (c) If relevant, consider translating estimates of relative risk into absolute risk for a meaningful time period | Not applicable |
| Other analyses | 17 | Report other analyses done—eg analyses of subgroups and interactions, and sensitivity analyses | Results |
| **Discussion** |  |  |  |
| Key results | 18 | Summarise key results with reference to study objectives | Discussion |
| Limitations | 19 | Discuss limitations of the study, taking into account sources of potential bias or imprecision. Discuss both direction and magnitude of any potential bias | Discussion |
| Interpretation | 20 | Give a cautious overall interpretation of results considering objectives, limitations, multiplicity of analyses, results from similar studies, and other relevant evidence | Discussion |
| Generalisability | 21 | Discuss the generalisability (external validity) of the study results | Discussion |
| **Other information** |  |  |  |
| Funding | 22 | Give the source of funding and the role of the funders for the present study and, if applicable, for the original study on which the present article is based | Abstract and Methods |

## Appendix 2: selection about the number of clusters

Figure 1 shows the Silhouette Coefficient and Elbow Coefficient (using within cluster sum of squares distance) computed for different numbers of clusters (k) from 2 to 12. One may choose a k of 2 because it yields the highest Silhouette Coefficient. However, when k=2, the Elbow Coefficient (aka sum of squared distances) is still very high. Thus, we sought to choose another point where the Silhouette Coefficient is still very high and also a low Elbow Coefficient. Therefore, we chose a k of 5 because it shows the 2nd highest Silhouette Coefficient, and the Elbow Coefficient is considerably low already (the criterion of reaching an elbow with the Elbow Coefficient is satisfied).

**Figure 1.**
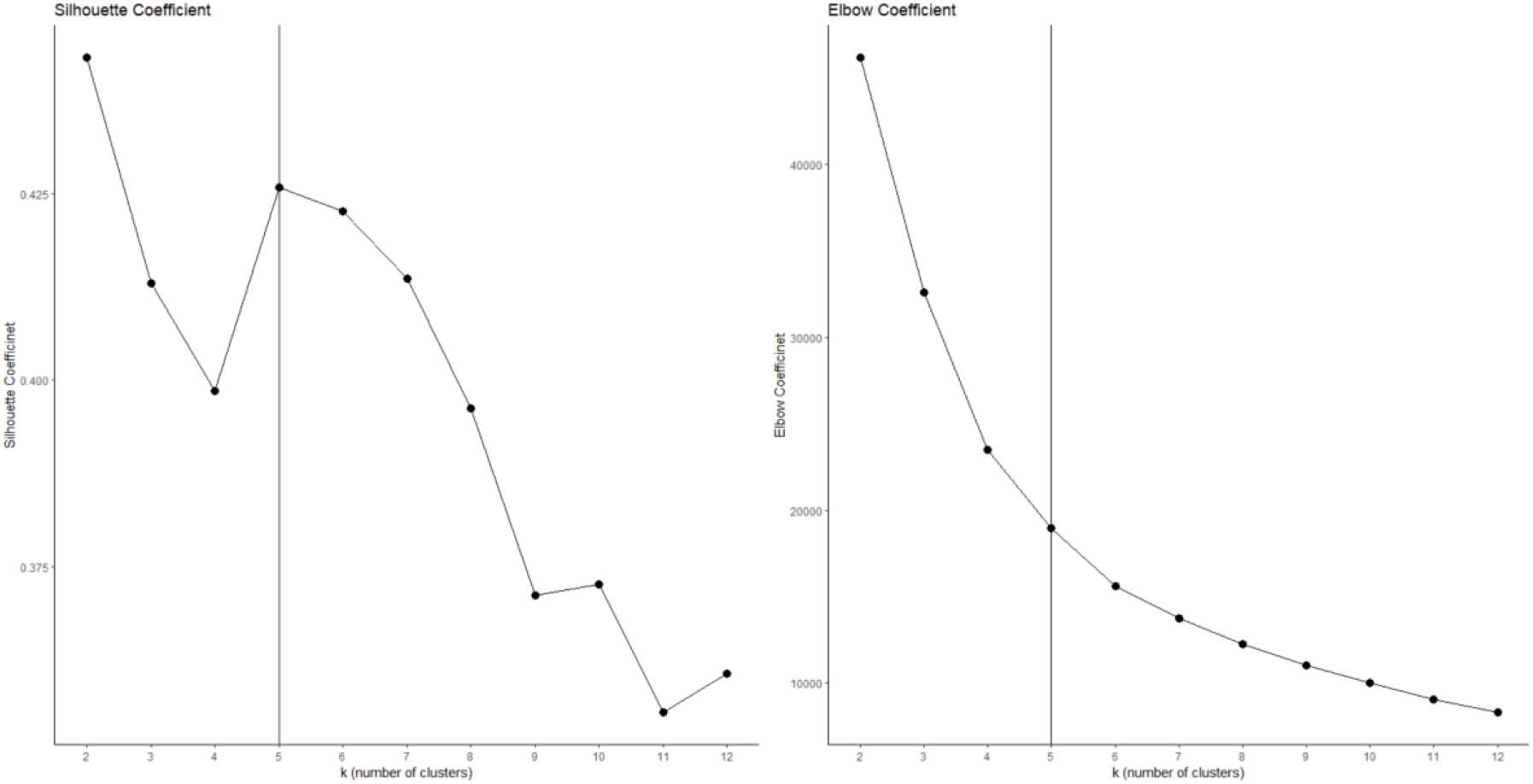
Silhouette Coefficient and Elbow Coefficient for different numbers of clusters.

## Appendix 3: Inter-clustering difference between the variable sex=female and Mortality7_ICU

**Figure 2.**
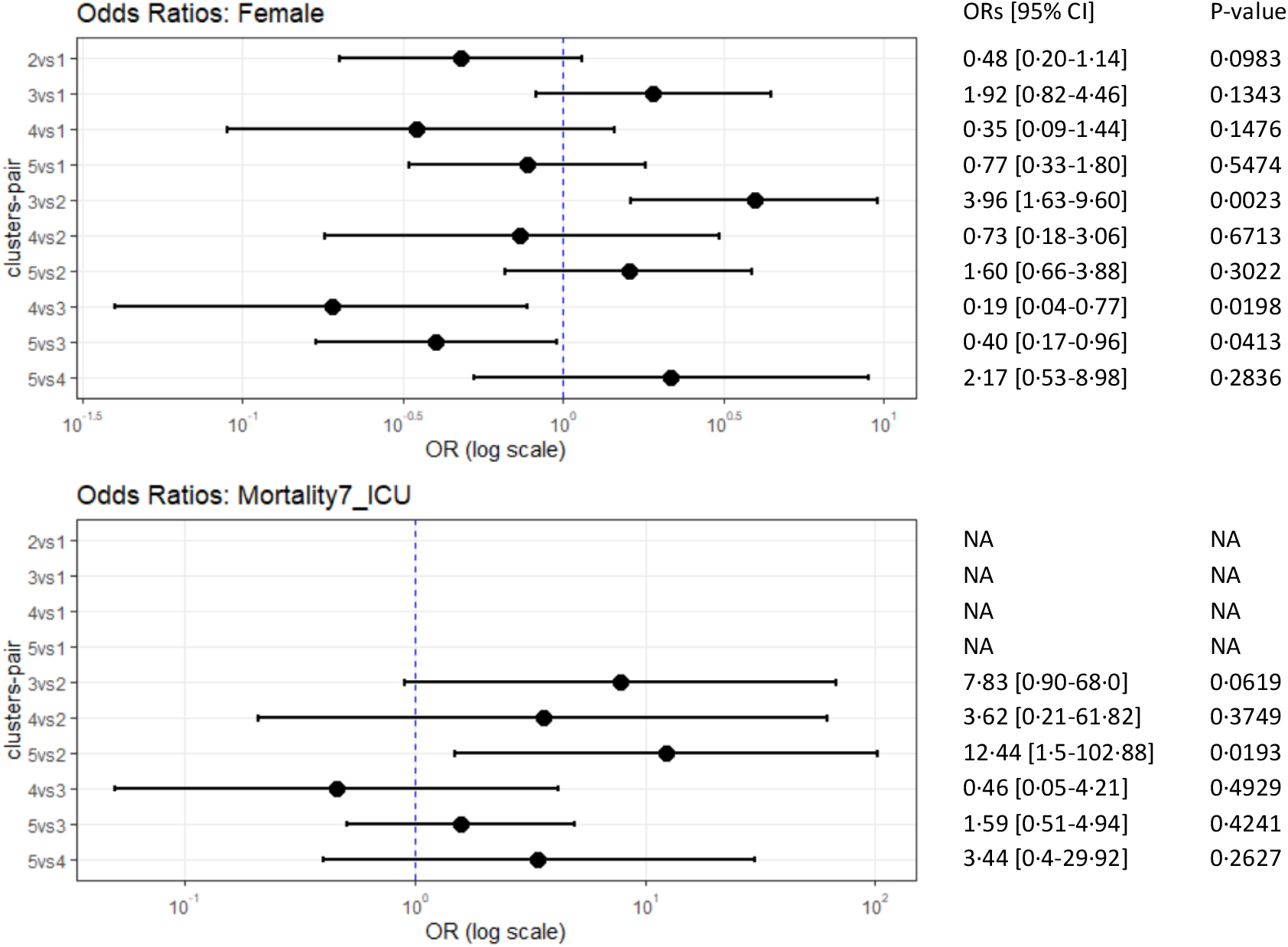
Odds Ratios with 95% CI and the corresponding p-value using Fisher’s exact test for two studied categorical variables: sex=Female, ICU mortality within 7. The Y-axis represents the clusters-pair that it compares. The X-axis represents the OR on the log-10 scale. Remark: there are OR values of NA in Mortality7_ICU, it’s because, in cluster 1, there were no patients who died within seven days after ICU admission.

## Notes

### Competing Interest Statement

The authors have declared no competing interest.

### Funding Statement

This study was funded by FONDO SUPERA COVID-19 by CRUE-Santander Bank grant SUBCOVERWD-19.

### Author Declarations

Ethics comittee of Universitat Politecnica de Valencia gave ethhical approval for this work. Ethics comittee of Hospital Clinic Universitari de Valencia gave ethhical approval for this work. Ethics comittee of Hospital Universitario 12 de Octubre gave ethhical approval for this work.

